# Critical success factors for high routine immunization performance: A multiple case study analysis of Nepal, Senegal, and Zambia

**DOI:** 10.1101/2022.11.08.22282076

**Authors:** Zoe Sakas, Kyra A. Hester, Anna S. Ellis, Emily Awino Ogutu, Katie Rodriguez, Robert A. Bednarczyk, Sameer Dixit, William Kilembe, Moussa Sarr, Matthew C. Freeman

**Affiliations:** Rollins School of Public Health, Emory University, Georgia, USA; Center for Molecular Dynamics Nepal, Kathmandu, Nepal; Center for Family Health Research in Zambia, Lusaka, Zambia; Institut de Recherche en Santé de Surveillance Epidemiologique et de Formation, Dakar, Senegal

## Abstract

**INTRODUCTION:** Vaccination averts an estimated 2-3 million deaths annually. Although vaccine coverage improvements across Africa and South Asia have remained relatively stagnant and below global targets, several countries have outperformed their peers with significant increases in routine immunization coverage. Examining these countries’ vaccination programs provides an opportunity to identify and describe critical success factors that may have supported these improvements.

**METHODS:** We selected three high-performing countries in regard to childhood vaccination: Nepal, Senegal, and Zambia. This multiple case study analysis was conducted using data from the Exemplars in Vaccine Delivery project within the Exemplars in Global Health program. We used qualitative analysis to investigate factors that contributed to high vaccination coverage through key informant interviews (KIIs) and focus group discussions (FGDs) at the national, regional, district, health facility, and community levels. We triangulated these findings with quantitative analyses using publicly available data, which are published elsewhere.

**RESULTS:** Our data revealed that the critical success factors for vaccine programming relied on the cultural, historical, and statutory context in which the interventions were delivered. In Nepal, Senegal, and Zambia, high immunization coverage was driven by 1) strong governance structures and healthy policy environments; 2) adjacent successes in health systems strengthening; 3) government-led community engagement initiatives; and 4) adaptation considering contextual factors at all levels of the health system.

**CONCLUSION:** Throughout the study, our analysis returned to the importance of defining and understanding the context, governance, financing, and health systems within a country, rather than focusing on any one intervention. This paper augments findings from existing literature by highlighting how contextual factors impact implementation decisions that have led to improvements in childhood vaccine delivery. Findings from this research may identify transferable lessons and support actionable recommendations to improve national immunization coverage in other settings.

**Key Messages:** *What is already known on this topic:* Immunization is a cost effective and life-saving public health intervention. The essential components of an effective vaccine delivery system are well-established, along with the behaviors related to routine immunization.

*What this study adds:* This study highlights how structural and contextual factors impacted the implementation of childhood vaccination programs in several countries with high vaccination coverage. By applying a positive deviant approach, we identify and describe drivers of immunization coverage that programmers and policy makers may utilize to better understand underlying factors within the system.

*How this study might affect research, practice, or policy:* Through focusing on countries with high routine immunization coverage, we examined how vaccine delivery systems may leverage components of existing governance structures and health systems to accelerate and sustain coverage. Operational definitions for governance, health systems strengthening, community engagement, and adaptive capacity, along with descriptions of how these processes were implemented in high-performing countries, may help other countries implement similar improvements.

## 1. Introduction

Vaccination is recognized as one of the most influential public health interventions, averting an estimated 2-3 million deaths annually [1–4]. The Global Vaccine Action Plan (GVAP) targeted at least 90% country-level coverage of the third dose of diphtheria, tetanus, pertussis vaccine (DTP3) among 1-year-old children [5], a globally recognized proxy for vaccination system performance [6]. However, by 2018 only 95 of the 193 World Health Organization (WHO) Member States achieved national GVAP targets, and less than one third met district level targets of 80% DTP3 [7–9].

In 2019, the African Region reported the lowest DTP3 coverage at 74%, a slight improvement from 71% coverage in 2010 [8]. DTP3 coverage in the South-East Asia Region substantially improved by 2019, to 91%; however, from 2000 to 2016, this region reported one of the lowest coverage rates with an average of 78% [8]. Within both regions, several countries outperformed their peers with significant increases in routine immunization coverage since 2000 [10]. Examining these countries’ vaccination programs provides an opportunity to identify and describe critical success factors that may have supported these improvements.

The essential components of an effective vaccine delivery system are well-established and include strong governance and leadership, healthcare financing, human resources, a robust supply chain, community engagement, and information systems for evidence-based decision-making [11–13]. Research on behaviors related to routine immunization focuses on recognized determinants of coverage, including intent to vaccinate, community access, and health facility readiness [14]. This paper augments findings from existing literature by highlighting how structural and contextual factors impact implementation decisions that have led to improvements in childhood vaccine delivery.

Our research describes how countries with high vaccination rates took similar paths to success [15–18]. Through investigating drivers of high routine immunization coverage, our findings revealed the importance of structural factors, including governance processes and collaboration between in-country stakeholders, which more broadly align with requirements for strong national health systems. Global DTP3 coverage has plateaued since 2010 at 86% [19]; identifying factors and mechanisms for accelerating progress is critical for expanding vaccination program reach and retention [9].The purpose of this study was to identify critical success factors that contributed to catalytic growth in childhood routine immunization coverage. Findings from this research may identify transferable lessons and support actionable recommendations to improve national immunization coverage in other settings [20].

## 2. Methods

This multiple case study analysis was conducted using data from the Exemplars in Vaccine Delivery project within the Exemplars in Global Health program [10, 20, 21]. We used qualitative analysis to investigate factors that contributed to high vaccination coverage through key informant interviews (KIIs) and focus group discussions (FGDs) at the national, regional, district, health facility, and community levels. We triangulated these findings with quantitative analyses using publicly available data, which are published elsewhere and referenced throughout this paper [10, 21].

Prior to data collection, we developed a conceptual model (Figure 1) to organize factors that impact childhood vaccine coverage globally. This model was based on the work of Phillips et al. and LaFond et al. alongside a broader, a priori, review of the vaccine literature [14, 22].

**Figure 1.**
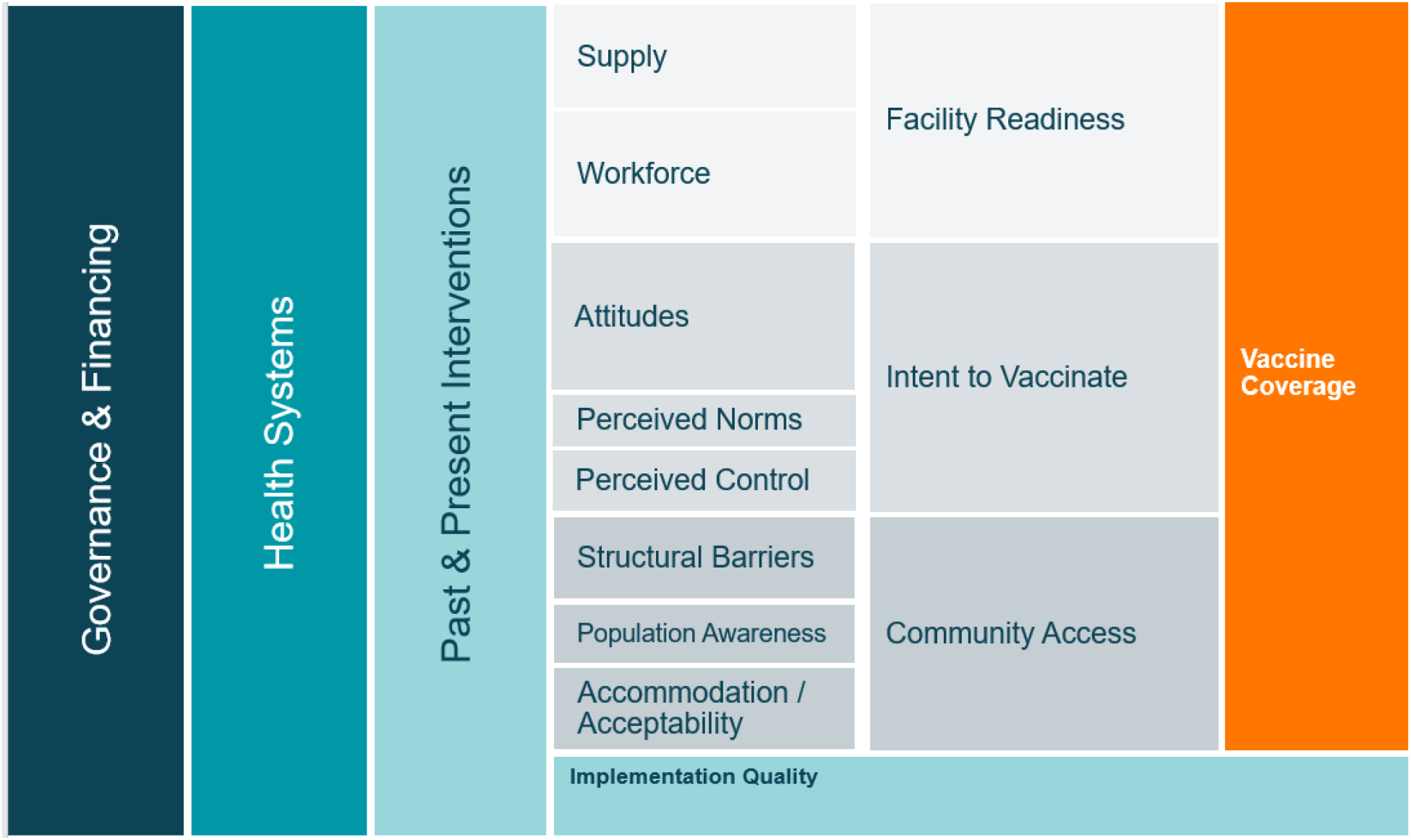
A priori conceptual model of the drivers of vaccine coverage.

### 2.1. Study sites

Methods for country, region, and district selection are detailed elsewhere [10, 15–17]. Briefly, countries were selected based on DTP1 and DTP3 coverage estimates, which served as proxies of the vaccine delivery system, with DTP1 as a proxy of access and DTP3 as a proxy of continued utilization of immunization services (Appendix 1) [10, 23, 24]. Three regions within each country were identified in consultation with national stakeholders and available data (Table 1). Partner organizations and Ministry of Health (MoH)^1^ officials facilitated site selection and data collection activities.

**Table 1:**
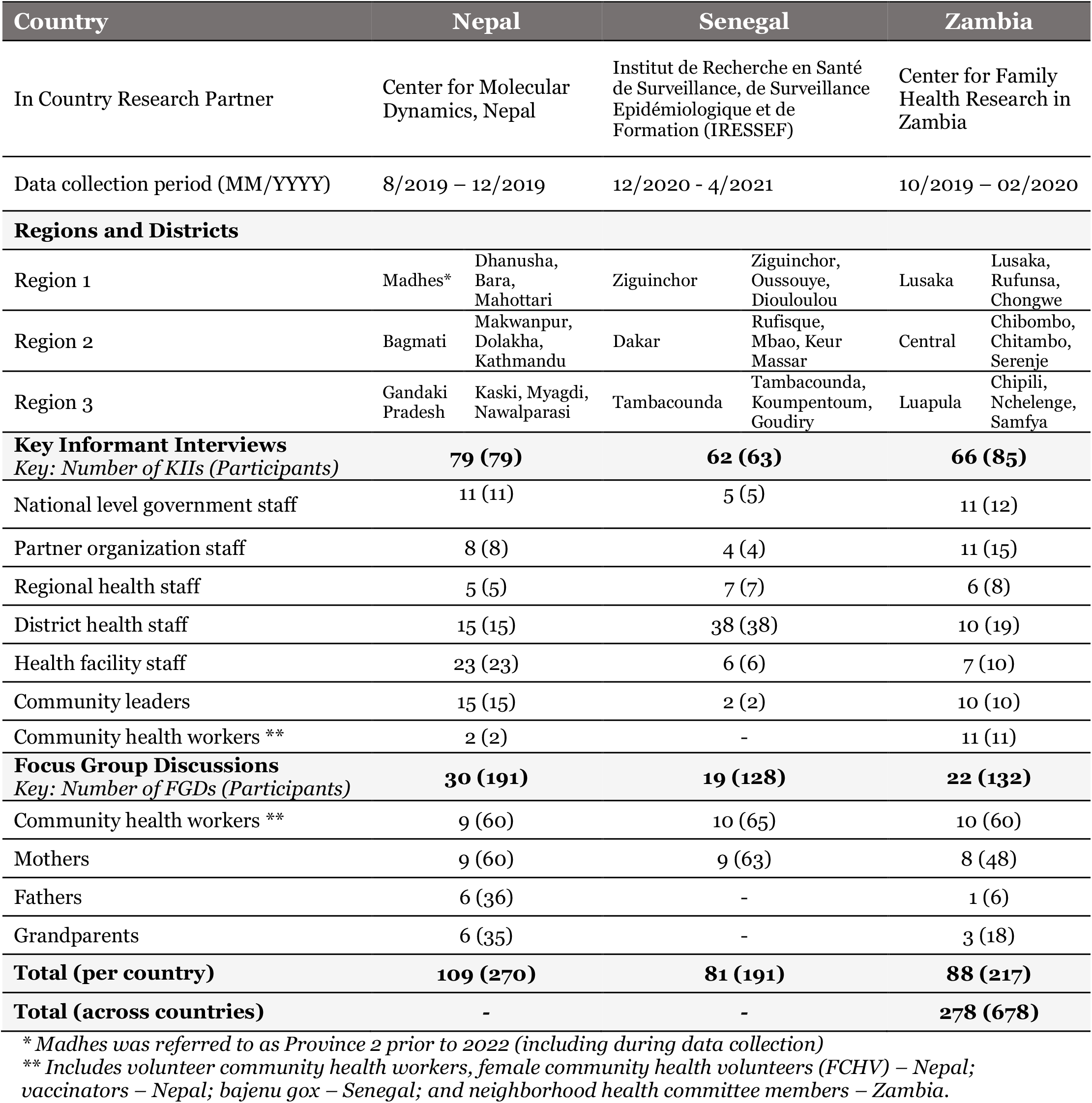
Summary of countries, regions, districts selected for research, and data collection activities.

### 2.2. Qualitative data collection and analysis

Qualitative data were collected between October 2019 and April 2021 at the national, regional, district, health facility, and community levels in Nepal, Senegal, and Zambia (Table 1). The interview guides were informed by the Consolidated Framework for Implementation Research (CFIR) [25] and the Context and Implementation of Complex Interventions (CICI) framework [26]. KII and FGD guides were translated into local languages by research assistants from the respective countries. All interview guides were piloted before use and adjusted iteratively throughout data collection. An initial list of KIIs was developed with local research partners and MoH officials with snowball sampling used to identify additional key informants. Our sampling approach aimed to include a diverse sample of participants in regard to geographic location and demographic qualities. Caregivers and volunteer community health workers were recruited for FGDs from health facility catchment areas with the assistance of local health staff. The duration of KIIs and FGDs averaged one and a half hours. KIIs and FGDs were audio-recorded with the permission of participants. Research files, recordings, and transcriptions were de-identified and password protected. We applied a theory-informed thematic analysis of the transcripts to identify critical success factors. Transcripts were coded and analyzed using MaxQDA2020 software (Berlin, Germany). We considered setting and participant roles while identifying key points and further contextualized data using historical documents and an a priori literature review. Qualitative data collection and analysis tools, including topic guides and codebooks, can be found on our Open Sciences Framework (OSF) page [27].

### 2.3. Supplemental quantitative data analysis

Quantitative data were collected through secondary datasets, which included information from the MOH in Zambia, Nepal, and Senegal among other partners. Data were used to estimate routine immunization coverage from 2000 to 2019 and to uncover trends related to catalytic improvements. Additional analyses were conducted to identify indicators that may be associated with immunization coverage success among low- and lower-middle-income countries using regression models to statistically test financial, development, demographic, and other country-level indicators. These quantitative analyses were published elsewhere. Findings from the quantitative analyses provided additional context for the qualitative findings which informed the results presented here.

### 2.4. Ethical approval

This study was considered exempt by the Institutional Review Board committee of Emory University, Atlanta, Georgia, USA (IRB00111474); approved by the Nepal Health Research Council (NHRC; Reg. no. 347/2019) in Kathmandu, Nepal; the National Ethical Committee for Health Research (CERNS; Comité National d’Ethique pour la Recherche en Santé) in Dakar, Senegal (00000174); the University of Zambia Biomedical Research Ethics Committee (Federal Assurance No. FWA00000338, REF. No. 166-2019); and the National Health Research Authority in Zambia. All ethics committees abide by the principles of the Declaration of Helsinki; all participants provided written consent.

### 2.5. Public involvement

The study team ensured the involvement of stakeholders, researchers, and policy makers in the global immunization sector during the design, implementation, and dissemination of this project. We utilized our Technical Advisory Group (TAG) to develop an initial set of questions for scoping visits to our exemplar countries, wherein study members met with in-country partners and experts to further refine the direction and nature of the questions. Scoping visit findings were used to create the final data collection tools [10]. As this is a hypothesis generating study, and no intervention was provided, data collection was targeted to conform to general standards of burden. We adhered to standard best-practices for qualitative research, iteratively implementing feedback from experts and the TAG. While there was community involvement (FGDs), the primary focus of data collection was centered on historical stakeholders who played key roles in the immunization program [10]. Upon completion of each country case study, findings were disseminated to in-country experts and country governments, along with the TAG, allowing for the study team to ground-truth results. Reports, and manuscripts, of findings were sent to those involved.

### 2.6 Sex and Gender Considerations

This study was hypothesis generating, and did not provide an intervention; regardless, the research team took gender differences into consideration. For our FGDs, we worked with groups of both mothers and fathers; fathers were included as to not fall into the bias of children’s health being a ‘woman’s issue’. FGDs were separated by gender to allow participants to feel comfortable sharing their thoughts.

Due to the historical nature of this study, our KII sample of key stakeholders and leaders leaned male due to prior gender biases and the tendency to place men in positions of importance or power. However, present-day positions were often filled by women, and we sampled women for KIIs as appropriate.

## 3. Results

### 3.1 Drivers of successful vaccination programs in exemplar countries

Following analysis of data from Nepal, Zambia, and Senegal, we revisited the conceptual framework developed during study conception (Figure 1). Although existing literature elaborates on factors related to intent to vaccinate, facility readiness, and community access, there is a gap in knowledge related to *how* policies and programs are operationalized to address these determinants – which relies heavily on the existing governance and health systems, engagement of community members (or end-users), and ability to adapt to the context to improve implementation. Our empirical data from Nepal, Senegal, and Zambia revealed that the critical success factors could be best understood by detailing the cultural, historical, and statutory context in which the interventions were delivered. This is in contrast to any specific modification or adaptation of key interventions or policies. We revised the initial conceptual framework based on findings from empirical data to illustrate a greater focus on these emergent factors (Figure 2). Definitions for the key domains can be found in Table 2.

**Table 2:**
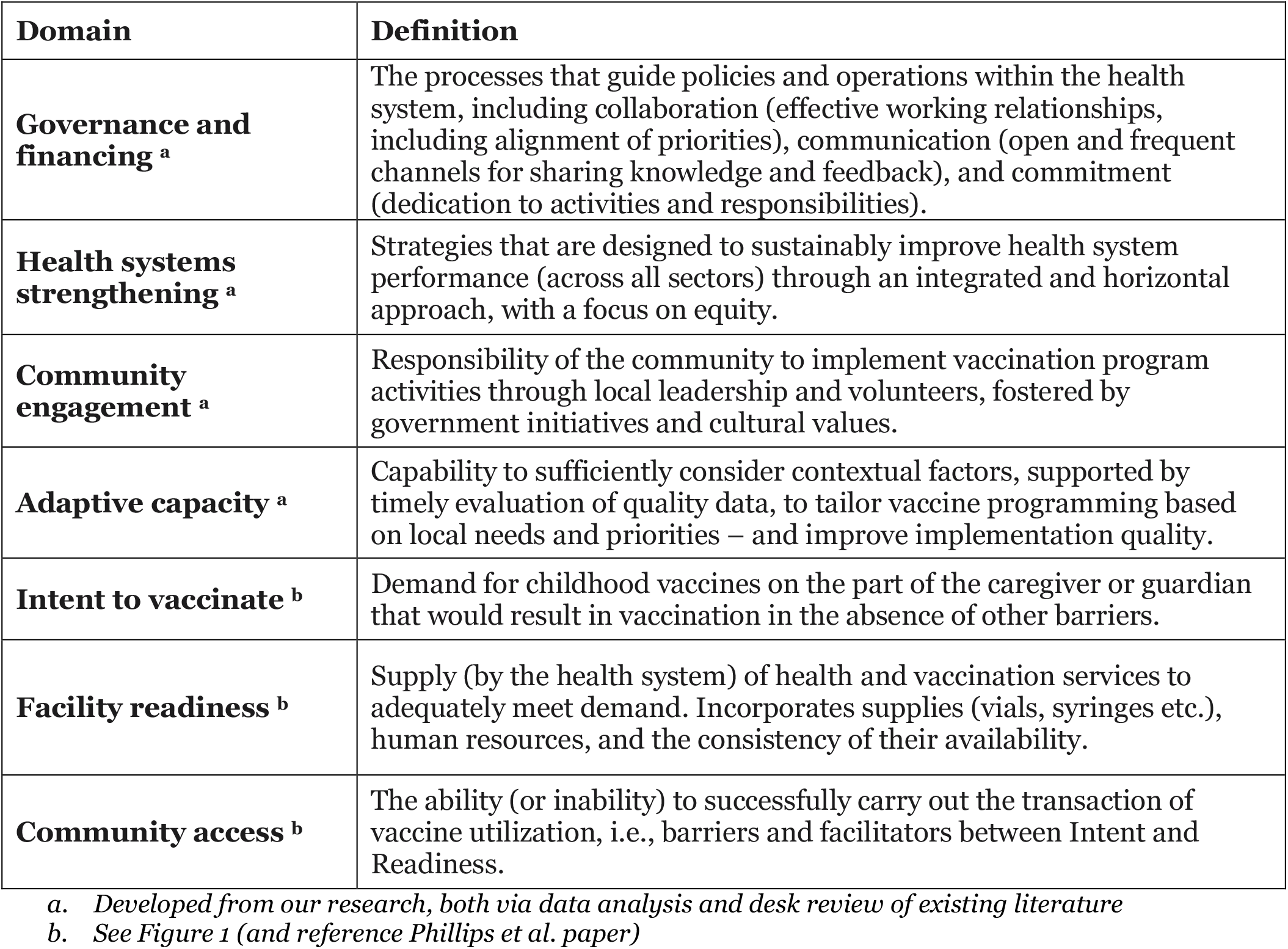
Definitions for drivers of vaccine coverage.

**Figure 2.**
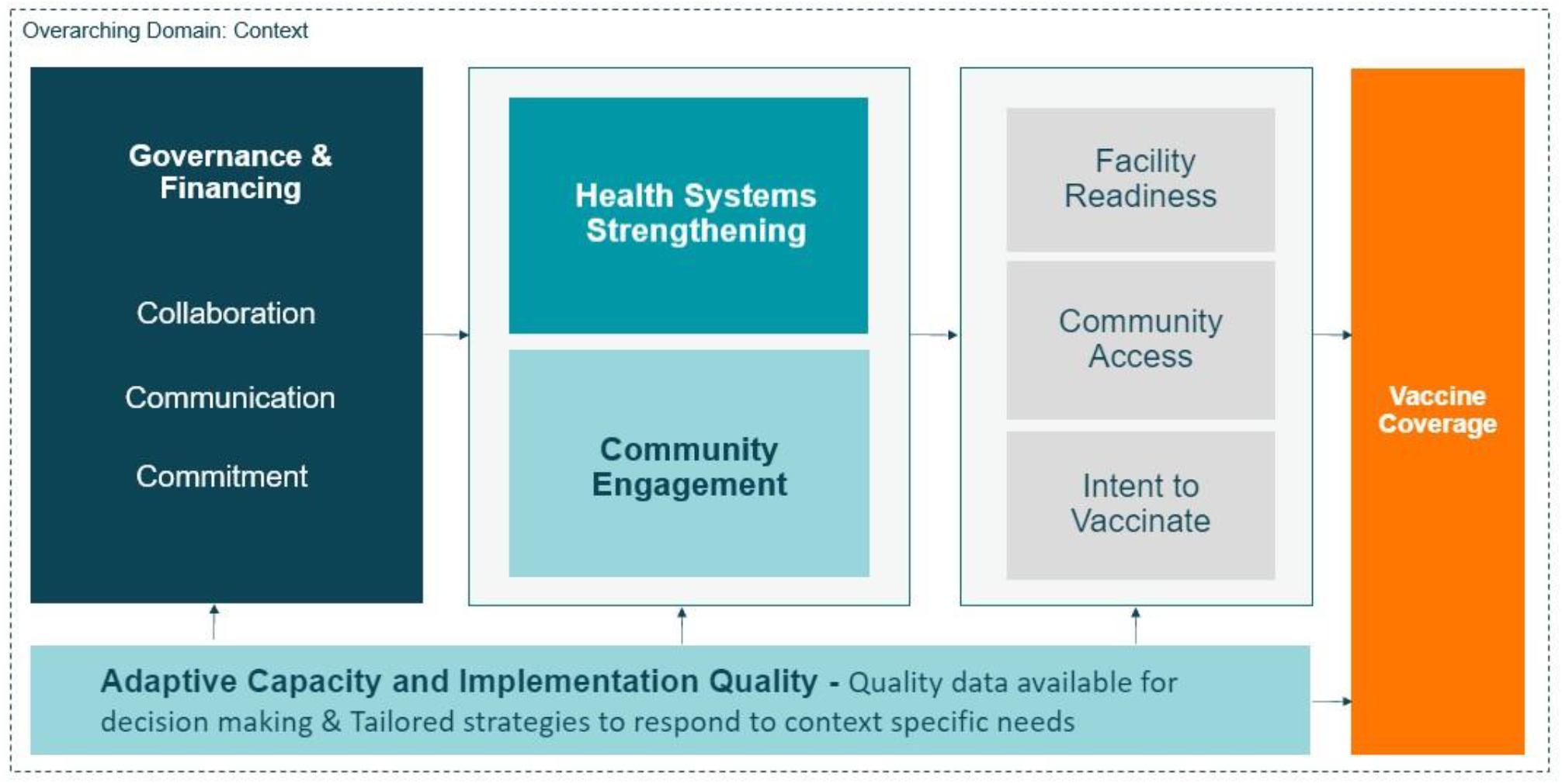
Revised conceptual framework for drivers of vaccine coverage.

### 3.2. Operationalization of successful vaccine delivery programs

In Nepal, Senegal, and Zambia, high immunization coverage was driven by 1) strong governance structures and healthy policy environments; 2) adjacent successes in health systems strengthening; 3) government-led community engagement initiatives; and 4) adaptation considering contextual factors at all levels of the health system. Below, we summarize similarities apparent in all three high-performing countries. Appendix 2 includes informative quotes as they relate to the four domains. More information on how policies and programs were implemented and adapted to local context is available elsewhere; quotes from each country and domain can be found in Appendix 2 [15–17].

### Strong governance structures and a healthy policy environment

Effective collaboration, open communication, and commitment to health and vaccines supported policy development and implementation of the vaccination programs in Nepal, Senegal, and Zambia.

Collaboration and communication enabled alignment of priorities, division of responsibilities, and coordination between levels. In all three countries, collaboration was apparent between external partners and the government, and between different levels of the health system. National fora were established for frequent and multi-disciplinary communication, including Inter-Agency Coordinating Committees (ICC) [28]. Regional, district, and community level reflection meetings were held to share and review data and gather feedback for strategic planning. Formal channels for supportive supervision and feedback, both top-down and bottom-up, supported service delivery improvements and reporting.

In all three countries, vaccination was a “priority” or “flagship” program recognized by the MOH. Prioritization of vaccination within the health sector supported sufficient resource allocation. In Nepal and Senegal, dedication to the health sector more broadly was codified through defining health as a right in their national constitutions. Commitment to health and vaccines underpinned policy development and enabled consistent service delivery. Community members could expect reliable and equitable access to vaccines due to the prioritization of activities by health personnel and the dedication of volunteer community health workers who supported outreach activities and spearheaded demand generation and educational campaigns.

> *“A provision was made in the constitution, and [vaccines] became a fundamental right, so the government has a responsibility to provide. And management of currently running programs also [focuses on] children who have missed vaccines*.*” (Nepal Health Research Council Staff, Nepal)*

### Adjacent successes in health systems strengthening

Improvements in the broader health systems of Nepal, Senegal, and Zambia – including health post expansion, training of volunteer community health workers, general capacity building, and prioritization of equitable health services – supported high immunization coverage.

Health post expansion and capacity building at subnational and community levels were mentioned by most key informants in all three countries as essential to improvements in vaccination coverage. Building health facilities in rural areas especially allowed parents to easily vaccinate their children, without having to travel far distances or rely on outreach. Improvements in vaccine service delivery also relied on improvements in general infrastructure including roads, electricity, and connectivity required for transportation, health facility operations, and data reporting. In all three countries, general health systems strengthening was centered around equity as a priority for decision making. Health post expansion and community health worker programs were strengthened to prioritize service delivery in hard-to-reach communities, minority populations, and remote areas.

> *“Previously, there was no public health department at a grassroots level, it was just at the National level… So, now the Public Health Specialist is on the ground to try to see that this primary health care activity is promoted. And I think with that supervision, with the introduction of the public health nurses, Health promotion departments, I think these interventions couldn’t be achieved higher in terms of immunization activities*.*” (District Health Director, Zambia)*

Ongoing efforts to address equity concerns include: 1) Overcoming geographical barriers through health post expansion and outreach services supported by national-level policy and external partner funding; and 2) Addressing social, economic, and cultural factors through tailored messaging and local ownership of vaccination programming.

Integrated decision making across health and social sectors, through a horizonal rather than vertical approach, facilitated effective allocation of resources (e.g., monetary, human, technical, educational) could be efficiently allocated across programs. For example, national fora and committees in all three countries included representatives from the immunization sector, maternal and child health, education and development, and financial offices. During meetings, sectors would align long-term priorities and discuss how to reach overall goals in terms of health and social outcomes.

### Government-led community engagement initiatives

Community ownership of vaccination activities and corresponding health outcomes was apparent in Nepal, Senegal, and Zambia, and even more so in high-performing districts within these countries. Responsibility of the community to implement vaccination programming through local leadership and volunteers was fostered by government initiatives and cultural values. National policies focused on involvement of traditional leaders and all national-level stakeholders reported on the importance of community actors. In some communities, female volunteers would work beyond their mandates to ensure all children were vaccinated – a decision motivated by dedication to their communities at large, empathy for the children in their neighborhoods, and a vague understanding of herd immunity.

Key informants from the national to community level reported that demand for vaccines was critical for improving routine immunization coverage. In addition to the work of volunteer community health workers, demand generation and public awareness were accomplished through national policies, media engagement, involvement of community actors, and school outreach activities.

> *“Community health workers play a very important role – they are in direct contact with the population. They are the ones who live in the community, and they identify more with the community. So, there is a relationship of trust between them and the population*.*” (National level government stakeholder, Senegal)*

Further analysis of community health worker programs and demand generation activities in Nepal, Senegal, and Zambia will be detailed in our upcoming publications [29].

### Adaptation at all levels of the health system

In Nepal, Senegal, and Zambia, decision-makers and implementers at the national, subnational, and community levels demonstrated their ability to consider contextual factors, evaluate or reflect on quality data, and tailor vaccine programming based on local needs and priorities.

Quality data were available for evidence-based decision making in all three countries. Data software was implemented at the district level in all countries - or in the case of Senegal, at the health post level - allowing stakeholders at all levels to utilize accurate and up-to-date coverage estimates. Lack of quality data in some areas may have hindered this process. However, when data were not available, community actors would adapt programming based on head counts conducted by volunteers and an anecdotal understanding of the contextual and cultural needs in their communities.

> *“We have been advising facilities that they need to own the data; they have to make sure that they utilize it by analyzing*… *If they are not meeting their targets, they need to reflect on the data and see where they are going wrong… For those [health facilities] that are performing well, we find out what they are doing, and we try to replicate to the other facilities that are not doing as well*.*” (District Health Officer, Zambia)*

Adapting strategies to respond to context-specific needs was essential for reaching children. In all three countries, flexible vaccine delivery days and outreach services supported unique scheduling needs (e.g., in Senegal, vaccinating children in the late afternoon during farming season). Outreach strategies were also tailored to accommodate contextual factors and address barriers to access. Understanding cultural and social values enabled community actors to tailor demand generation activities so that community members would be more apt to adopt positive health behaviors. Some examples of this may be using a local language, involving religious or community leaders in engagement and sensitizing, or addressing cultural or social barriers directly through personal communication.

### 3.3. Interventions and programs across high-performing countries

Key informants discussed a myriad of interventions and programs that they believed led to the improved and sustained vaccination coverage in their countries (Table 3). Interventions are organized by implementation level; if an intervention was implemented at multiple levels of the health system, it may be repeated in the table. Although the specific components of vaccine programming were not identical across countries, many of the general themes overlapped (e.g., the scope of community-level health committees varied between countries, but the purpose/objective of these groups was similar).

**Table 3:**
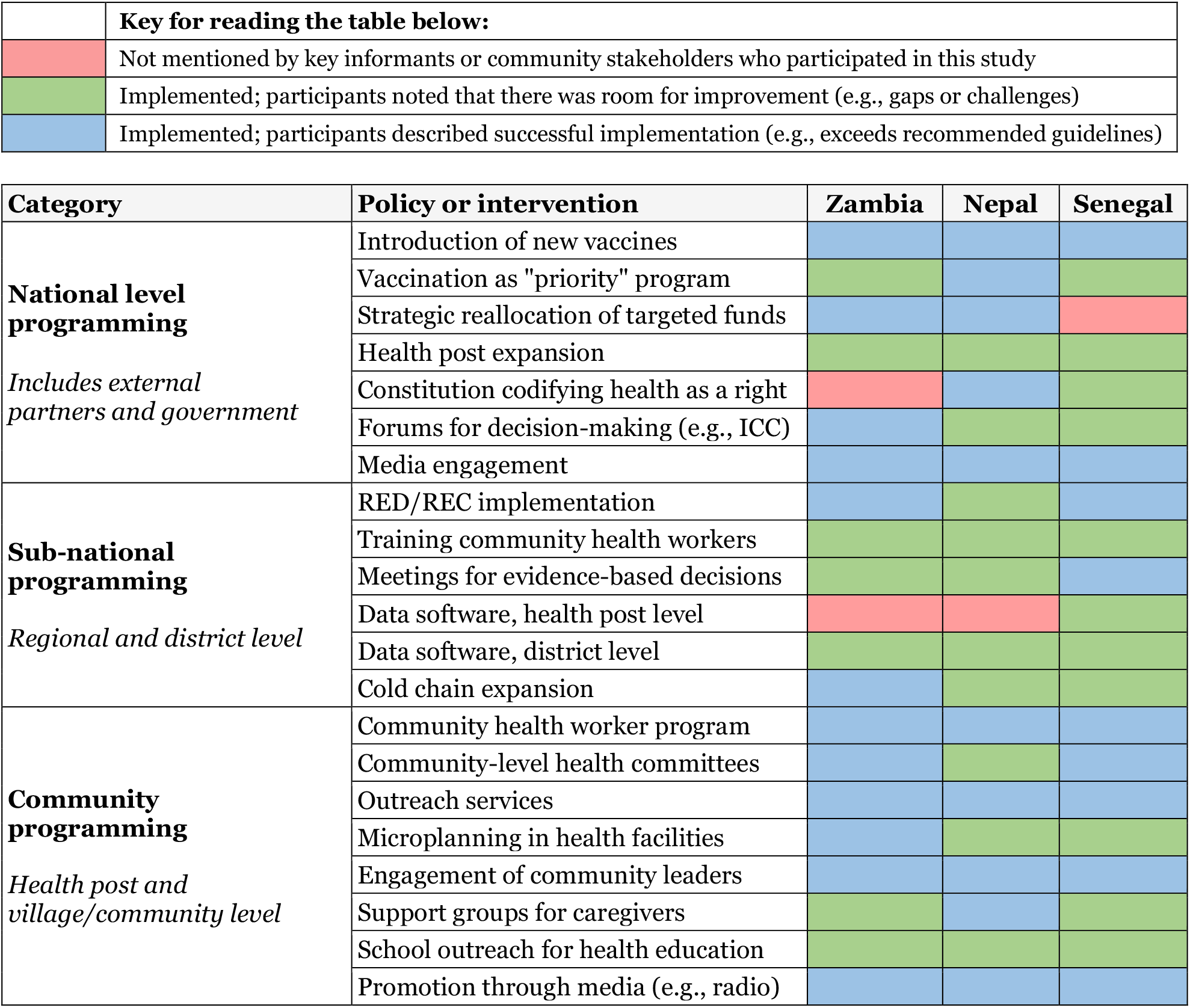
Key interventions and programs implemented to improve routine immunization coverage in Nepal, Senegal, and Zambia from 2000 to 2019, based on KII and FGD data.

## 4. Discussion

Notions of strengthening national governance structures have historically been recognized through country-level application of global guidelines to improve vaccine delivery and health systems; however, existing research lacks concrete descriptions of what this entails. This paper augments findings from existing literature by highlighting how structural and contextual factors impact implementation decisions that have led to improvements in childhood vaccine delivery. We developed evidence-derived operational definitions of structures and processes that we identified as critical to the success of vaccine delivery systems in countries with high routine immunization coverage. We described *how* these processes were utilized in high-performing countries to drive catalytic improvements in coverage. Previous research has not explored the details or importance of governance, financing, health systems, or other contextual factors; our original framework based on preexisting literature (Figure 1) was missing complexity in these components, which is indicative of the limited knowledge in this area. Describing how national governance structures operate with the consideration of contextual factors was essential to our understanding of how global guidelines should be adapted to reflect local priorities and challenges. Ignoring a structured approach to understanding these factors could lead to repeated instances of sub-optimal program performance.

Our data points to the need for programmers and policy makers to better understand underlying governance structures. The structures and processes we examined in Nepal, Senegal, and Zambia supported adaptive decision-making based on context-specific needs and priorities. Adaptation at all levels enabled tailored and targeted programming based on evaluation of high-quality data. While health system improvements led by national governments were essential, our data also highlighted the importance of a deliberate and consistent focus on community engagement, which aligns with existing literature [1,27,28]. Community involvement in vaccine programming facilitated the government’s understanding of cultural and social barriers and enablers which contributed to effective adaptation of interventions to increase coverage. Community ownership was fostered through national policies and resources for capacity building, engagement of traditional leaders, and the strength of the community health worker programs.

We found that many improvements in immunization coverage emerged from adjacent successes in health systems strengthening (HSS). While an unsurprising finding that health system strengthening would be an essential ingredient in support of vaccine system strengthening, it points to the need for a more integrated, rather than siloed, approach to child health. HSS has become a priority in recent decades as disruptions due to natural disasters, disease outbreaks, or civil unrest have exposed weaknesses in health systems [30, 31]. Although we focused on examining success factors for vaccine delivery, our data also suggests that there are still gaps and challenges in exemplar countries, including: 1) equitable access (e.g., lack of health posts in rural and remote areas); 2) long-term financing (e.g., most funding comes from external partners); 3) poor infrastructure (e.g., roads, electricity); and 4) a heavy reliance on volunteer community health workers which experienced vacancies and decreased morale in some cases.

In Nepal, Senegal, and Zambia, overall commitment to health and vaccines strengthened governance processes and supported vaccination efforts through prioritization of activities and resource allocation. Following the completion of GVAP in 2020, stakeholders reported that lack of consideration of existing structures and contextual factors limited the plan’s success [32]. In addition, the monitoring and evaluation processes proposed through GVAP were often seen as unrealistic [9, 32, 33]. Suggested improvements included engaging with countries to accurately and sufficiently consider local context while setting global goals and targets and promoting country ownership of strategic planning [32, 33]. Our research revealed that national government ownership of strategic planning was critical to these countries’ success, which aligns with the critical reflection of GVAP and other existing literature.

Overall, the strategies implemented by exemplar countries align with the WHO Comprehensive Framework of Strategies and Practices for Routine Immunization, which focus on managing the immunization program (e.g., political commitment, partnerships), mobilizing communities, generating demand, and monitoring progress [1]. Throughout the study, our analysis returned to the importance of defining and understanding the context, governance, financing, and health systems within a country, rather than focusing on any one intervention. This highlights the importance of further research into these structural factors. We examined how these strategies were operationalized to improve coverage in Nepal, Senegal, and Zambia, and how they can be applied in other settings. Understanding how governance processes function in high-performing countries may allow others to build stronger systems, adapt guidelines, and reflect on progress made in the field. Our research introduces areas of inquiry for further investigation, as the critical success factors that primarily emerged in all three exemplar countries are not the current focus within existing literature around determinants for vaccine coverage. as the critical success factors that primarily emerged in all three exemplar countries are not the current focus within existing literature around determinants for vaccine coverage.

## 5. Limitations

This study has several limitations. First, it was difficult to determine causation because we focused on countries that were successful in vaccine delivery but were unable to carry out a similar analysis in a country with lower vaccination coverage to compare. Second, the research tools focused on the factors that drove catalytic change and did not focus on interventions or policies that were unsuccessful. Third, using qualitative methods to understand historical events was challenging; interviewees often spoke about current experiences rather than discussing historical factors. However, we probed respondents to reflect on longitudinal changes in the immunization program. Additional country-specific limitations can be found elsewhere [15–17].

## 6. Conclusion

Through focusing on countries with high routine immunization coverage, we examined how vaccine delivery systems may leverage components of existing governance structures and health systems to accelerate and sustain coverage. Operational definitions for governance, health systems strengthening, community engagement, and adaptive capacity, along with descriptions of how these processes were implemented in high-performing countries, may help other countries implement similar improvements. Our findings highlight the need for program managers and policy makers to understand and consider the strengths and limitations of existing structures while adapting to the emergent needs and priorities relevant to the context. Our study looked retroactively on how and why countries succeeded in achieving high early childhood immunization coverage. The COVID-19 pandemic has highlighted the fragility in our health systems, and specifically how robust vaccine systems can rise to the challenge of emergent vaccination needs. While our findings are not immediately applicable to the current COVID vaccination needs, our underlying approach to understand context remain relevant to consider as countries strive to vaccinate their populations in the ongoing pandemic [34].

## Data Availability

Data are not publicly available as all data are confidential. De-identified data may be available upon request.

https://osf.io/7ys4a/

## Acknowledgements

We thank the Center for Family Health Research in Zambia, the Center for Molecular Dynamics Nepal, and the Institut de Recherche en Santé de Surveillance Epidemiologique et de Formation in Dakar, Senegal. We gratefully acknowledge the participants who gave their time and insights to help us better understand the vaccine delivery systems of Zambia, Nepal, and Senegal, along with facilitators from their respective Ministries of Health.

In addition, we thank Sarah Chesemore, Anna Rapp, Tove Ryan, and Ethan Wong from the Bill and Melinda Gates Foundation; Kate Buellesbach, Nancy Fullman, Nathaniel Gerthe, Gloria Ikilezi, Caitlyn Mason, David Phillips, and Oliver Rothschild, Jordan-Tate Thomas, and Angela Wang from Gates Ventures; and the Vaccine Exemplars Research Advisory Group for their insights, specifically Agnes Binagwaho, Laura Craw, Carolina Danovaro, Anuradha Gupta, Heidi Larson, Penelope Masumbu, Kate O’Brien, Helen Rees, Lora Shimp, and Aaron Wallace.

## Funding

This work was supported by the Bill & Melinda Gates Foundation, Seattle, WA (OPP1195041) with a planning grant from Gates Ventures, LLC, Kirkland, WA.

## Declaration of competing interests

The authors declare that they have no known competing financial interests or personal relationships that could have appeared to influence the research reported in this paper.

## Footnotes

1 *Note that each country has a slightly different name for their Ministry of Health (e*.*g*., *Ministry of Health and Population in Nepal; Ministry of Health and Social Action in Senegal), but all will be referred to as “MoH” throughout for simplicity*.

## Notes

### Competing Interest Statement

The authors have declared no competing interest.

### Clinical Protocols

https://bmjopen.bmj.com/content/12/4/e058321

### Author Declarations

This study was considered exempt by the Institutional Review Board committee of Emory University, Atlanta, Georgia, USA (IRB00111474); approved by the Nepal Health Research Council (NHRC; Reg. no. 347/2019) in Kathmandu, Nepal; the National Ethical Committee for Health Research (CERNS; Comite National d Ethique pour la Recherche en Sante) in Dakar, Senegal (00000174); the University of Zambia Biomedical Research Ethics Committee (Federal Assurance No. FWA00000338, REF. No. 166-2019); and the National Health Research Authority in Zambia.

## References

1. World Health Organization, Immunization Agenda 2030: A Global Strategy to Leave No One Behind. 2020.

2. Vanderslott, S. and T. Marks, Charting mandatory childhood vaccination policies worldwide. Vaccine, 2021.

3. Gavi the Vaccine Alliance. Sustainable Development Goals. 2020 18 February, 2020; Available from: https://www.gavi.org/our-alliance/global-health-development/sustainable-development-goals.

4. Piot, P., et al., Immunization: vital progress, unfinished agenda. Nature, 2019. 575(7781): p. 119–129.

5. World Health Organization, Global Vaccine Action Plan 2011–2020 2013: USA.

6. World Health Organization and UNICEF, Global Immunization Vision and Strategy 2006-2015. 2005: New York, NY.

7. World Health Organization. Immunization coverage. 2020 July 15, 2020 August 4, 2020]; Available from: https://www.who.int/news-room/fact-sheets/detail/immunization-coverage.

8. World Health Organization and UNICEF, WHO-UNICEF estimates of DTP3 coverage. 2020.

9. World Health Organization, GVAP review and lessons learned: methodology, analysis and results of the stakeholder consultations, in Annex to the Global Vaccine Action Plan 2011-2020. 2019: Geneva.

10. Bednarczyk, R.A., et al., Protocol: Identification and evaluation of critical factors in achieving high and sustained childhood immunization coverage in selected low- and lower-middle income countries. medRxiv, 2021: p. 2021.12.01.21267018.

11. Shen, A.K., R. Fields, and M. McQuestion, The future of routine immunization in the developing world: challenges and opportunities. Global health, science and practice, 2014. 2(4): p. 381–394.

12. World Health Organization, Global Routine Immunization Strategies and Practices (GRISP): a companion document to the Global Vaccine Action Plan (GVAP). 2016, Geneva: World Health Organization.

13. World Health Organization, Reaching Every District (RED): A guide to increasing coverage and equity in all communities in the African Region. 2017.

14. Phillips, D.E., et al., Determinants of effective vaccine coverage in low and middle-income countries: a systematic review and interpretive synthesis. BMC health services research, 2017. 17(1): p. 681–681.

15. Rodriguez, K., et al., Critical success factors for routine immunization performance: A case study of Zambia 2000 to 2018. medRxiv, 2021: p. 2021.11.30.21267060.

16. Sakas, Z., et al., Critical success factors for high routine immunization performance: A case study of Senegal. medRxiv, 2022: p. 2022.01.25.22269847.

17. Hester, K.A., et al., Critical success factors for high routine immunization performance: A case study of Nepal. medRxiv, 2022: p. 2022.01.28.22270023.

18. Noor, A.M., Country ownership in global health. PLOS Global Public Health, 2022. 2(2): p. e0000113.

19. WHO and UNICEF, Immunization, DPT (% of children ages 12-23 months), W.a. UNICEF, Editor. 2022.

20. Carter, A., et al., A framework for identifying and learning from countries that demonstrated exemplary performance in improving health outcomes and systems. BMJ global health, 2020. 5(12): p. e002938.

21. Exemplars in Global Health. Making Better Decisions in Global Health: Understand Positive Outliers to Inform Policy and Practice. 2021; Available from: https://www.exemplars.health/.

22. LaFond, A., et al., Drivers of routine immunization coverage improvement in Africa: findings from district-level case studies. Health policy and planning, 2015. 30(3): p. 298–308.

23. World Health Organization and UNICEF, WHO-UNICEF estimates of national immunization coverage (WUENIC). 2020: Geneva, Switzerland.

24. Nepal Demographic Health Survey 2001, 2006, 2011, 2016, 2017, DHS Program, Editor.

25. Damschroder, L.J., et al., Fostering implementation of health services research findings into practice: a consolidated framework for advancing implementation science. Implementation science : IS, 2009. 4: p. 50–50.

26. Pfadenhauer, L.M., et al., Making sense of complexity in context and implementation: the Context and Implementation of Complex Interventions (CICI) framework. Implementation science : IS, 2017. 12(1): p. 21–21.

27. Hester K, Open Sciences Framework (OSF) page: Assessing sustainability factors for rural sanitation coverage following the SSH4A approach. https://osf.io/caz9b/?view_only=a1f16382d78b445ab34cc2afedca0cb0.

28. Sakas, Z., et al., The role of Zambia’s expansive ICC in supporting efficient, effective, and evidence-based vaccine and health sector programming. medRxiv, 2021: p. 2021.11.29.21267021.

29. Hester, K.A., et al., Critical interventions for demand generation in Zambia, Nepal, and Senegal with regards to the 5C psychological antecedents of vaccination. medRxiv, 2022: p. 2022.04.25.22274035.

30. Ozawa, S., L. Paina, and M. Qiu, Exploring pathways for building trust in vaccination and strengthening health system resilience. BMC health services research, 2016. 16(Suppl 7): p. 639–639.

31. World Health Organization Maximizing Positive Synergies Collaborative Group, An assessment of interactions between global health initiatives and country health systems. The Lancet, 2009. 373(9681): p. 2137–2169.

32. Hwang, A., et al., Global Vaccine Action Plan Lessons Learned II: Stakeholder Perspectives. Vaccine, 2020. 38(33): p. 5372–5378.

33. Cherian, T., et al., Global Vaccine Action Plan lessons learned III: Monitoring and evaluation/accountability framework. Vaccine, 2020. 38(33): p. 5379–5383.

34. Dixit, S.M., et al., Addressing disruptions in childhood routine immunisation services during the COVID-19 pandemic: perspectives from Nepal, Senegal and Liberia. BMJ Global Health, 2021. 6(7): p. e005031.

