## Appendix 1 for "Critical success factors for high routine immunization performance: A multiple case study analysis of Nepal, Senegal, and Zambia"

Appendix 1: Coverage data for Nepal, Zambia, and Senegal

**DHS DTP3 coverage in Zambia, by Province, 2000 – 2016**

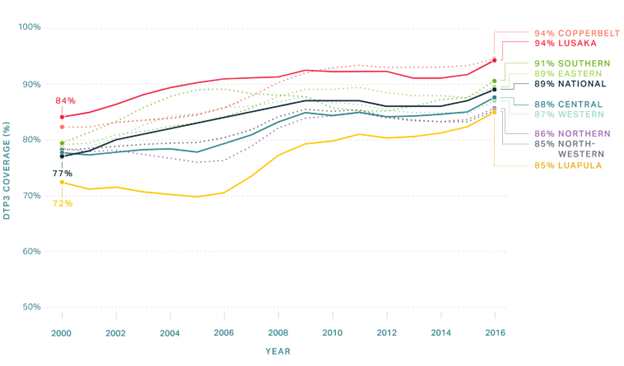

**DHS DTP3 coverage in Nepal, by Province, 2000 – 2016**

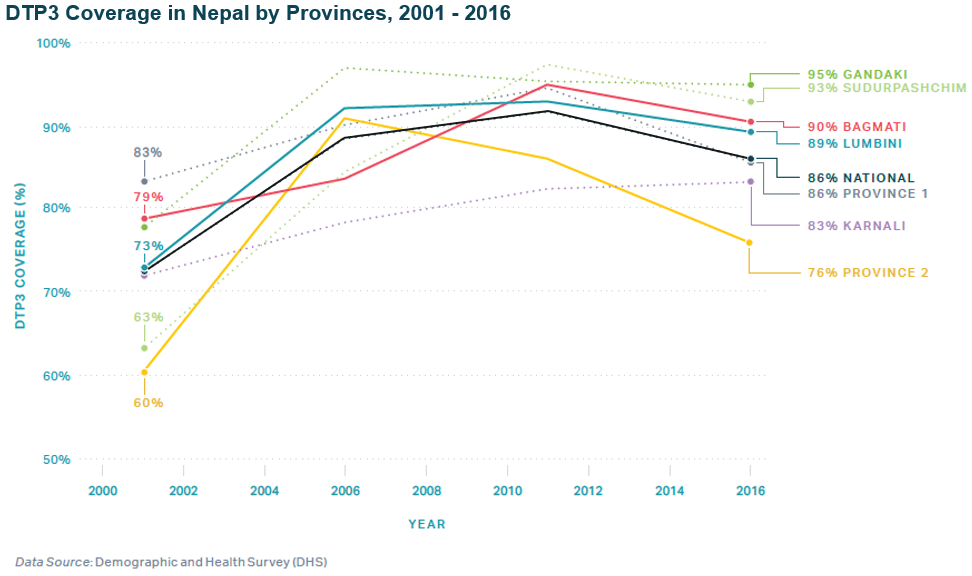

**DHS DTP3 coverage in Senegal, by Province, 2000 – 2016**

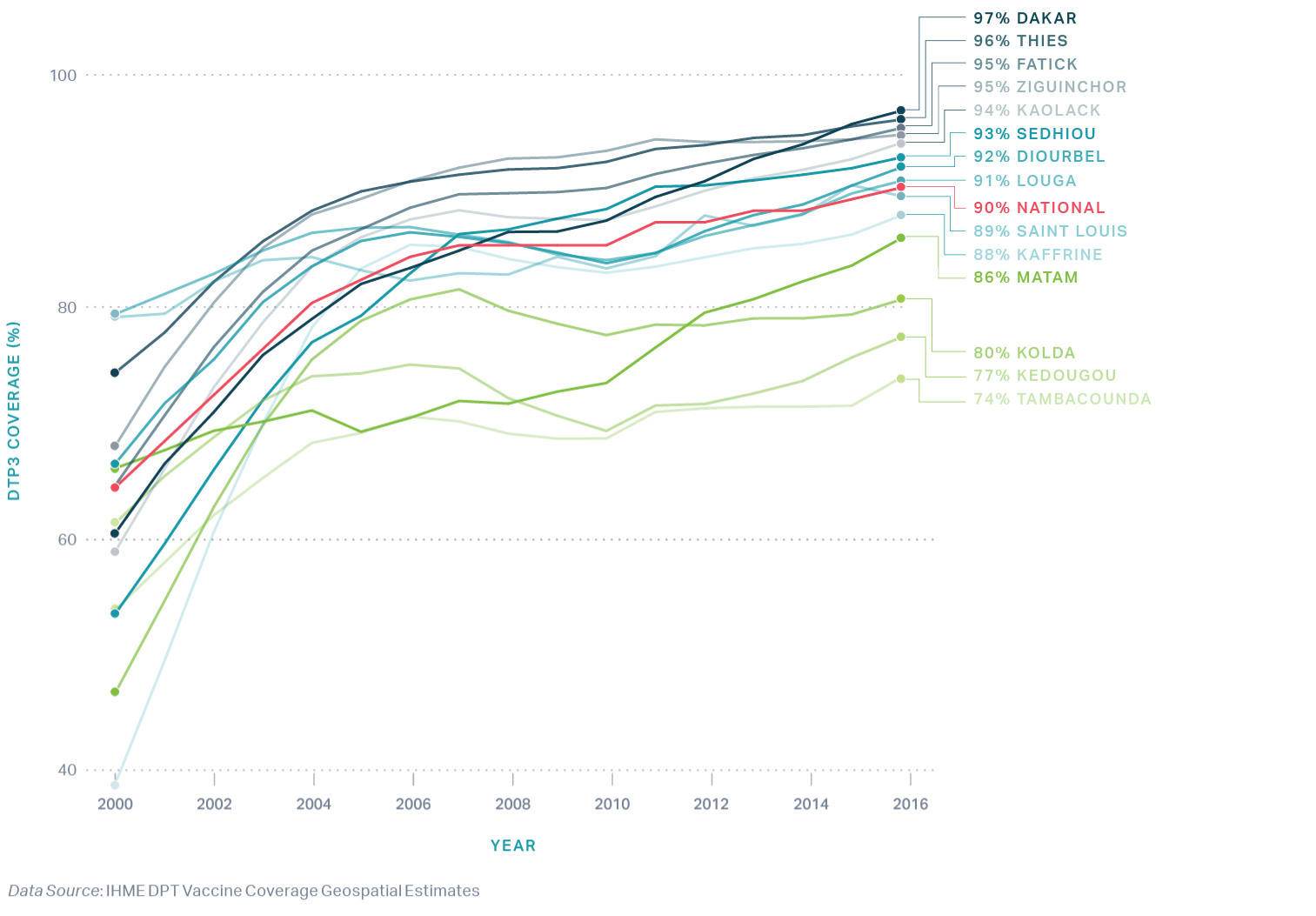

**DTP3 coverage from 2000 to 2018 of Nepal, Senegal, and Zambia**

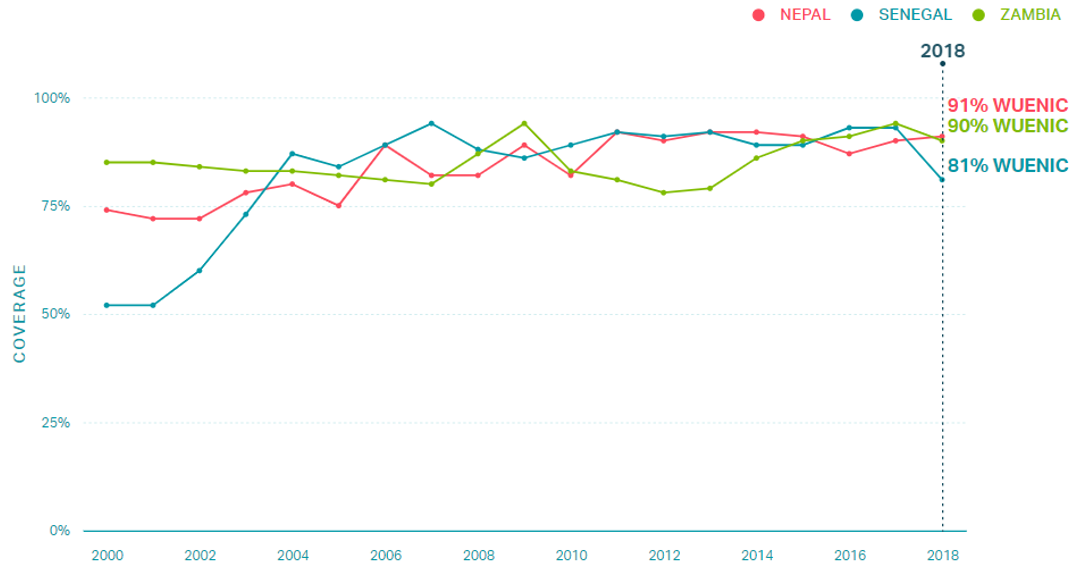
