## Appendix 2 for "Critical success factors for high routine immunization performance: A multiple case study analysis of Nepal, Senegal, and Zambia"

**Appendix 2: Additional key informant quotes, by country and domain, illustrating drivers for vaccine coverage from the conceptual framework [15-17]**

| **Domain** | **Nepal** | **Senegal** | **Zambia** |
| --- | --- | --- | --- |
| Governance and financing  The processes that guide policies and operations within the health system, including collaboration (effective working relationships, including alignment of priorities) and commitment (dedication to activities and responsibilities). | *“A provision was made in the constitution, and [vaccines] became a fundamental right, so the government has a responsibility to provide. And management of currently running programs also [focuses on] children who have missed vaccines.” (Nepal Health Research Council Staff)* | *“When we have a strategic document to work on, the partners get involved according to their area of expertise*, *and we work together when it comes to implementing and monitoring the vaccine program. In this teamwork, you cannot tell who is the partner or who is from the Ministry of Health, because we work hand in hand. We are in the same boat, we want to have success.” (External partner at the national level)* | *“Once you have identified that there is an issue that needs attention, these policies are revised and updated so that they match what is current. [Policies] are more or less like living documents to match the prevailing circumstances. Our planning has improved quite a lot because it’s now bottom-up. So, we are the ones that have a lot of input in these documents.” (Provincial Health Officer)* |
| Health systems strengthening  Strategies that are designed to sustainably improve health system performance (across all sectors) through an integrated and horizontal approach, with a focus on equity. | *“No wards will be deprived of health centers, and if the health center isn’t enough to provide health services, then village clinics, immunization centers (outreach clinics), and urban health clinics will be established. If services are provided from these health centers, quality of the vaccines are maintained, and health centers are accessible to all - then good immunization coverage can be achieved.” (Department of Health Services within the MOH)* | *“We tried to see what was hidden behind the weak coverage - especially the poor availability of resources including health structures and sufficient staff... a diagnosis was made to improve the indicators, health structures, i.e. creating a lot of health posts, and recruiting staff to be set up in each health post, or at least one vaccination unit to really try to reach out to the entire population in order to have good coverage.” (National level stakeholder)* | *“Previously, there was no public health department at a grassroots level, it was just at the National level… So, now the Public Health Specialist is on the ground to try to see that this primary health care activity is promoted. And I think with that supervision, with the introduction of the public health nurses, Health promotion departments, I think these interventions couldn’t be achieved higher in terms of immunization activities.” (District Health Director)* |
| Community engagement  Responsibility of the community to implement vaccination program activities through local leadership and volunteers, fostered by government initiatives and cultural values. | *“Mothers’ groups are responsible to give education and information to the parents.... If they don’t support the program, then it won't be successful. They are the ones who build awareness and give knowledge and information to the people at the grassroots level.” (Department of Health Services in the Ministry of Health and Population)* | *“Community health workers play a very important role – they are in direct contact with the population. They are the ones who live in the community, and they identify more with the community. So, there is a relationship of trust between them and the population.” (National level government stakeholder)* | *"We make sure that the community is fully engaged and aware of the benefit of ensuring that their children are vaccinated. It is important that we ensure that within the community, the key gatekeepers are helping us." (Provincial Health Officer)* |
| Adaptive capacity  Ability to sufficiently consider contextual factors, supported by timely evaluation of quality data, to tailor vaccine programming based on local needs and priorities. | *“Using HMIS, which is our biggest data source, we send daily, weekly, or monthly reports to districts using phones. We visit districts and contact the responsible person. We understand the situation and we try to solve those problems. We don’t depend on the system. We explore, analyze, and address if there are issues outside the system. We do review meetings at the end of the year from the lessons we learned. We also make periodic updates.” (Regional MoH Director)* | *“During regional-level meetings, we hear good ideas from the districts. When Dakar had a measles epidemic, we brought together all the districts to tell us how to plan in order to have good measles vaccine coverage. Some of the districts came up with strategies that we found very inspiring. One of them was to hold the vaccination sessions beyond regular work hours to reach the people who are not free during the day. The other strategy was to organize sessions on weekends. As soon as we put these strategies in place, we noticed that the coverage increased right away. It dawned on us that the parents did not refuse the vaccination but that they were busy by their daily occupations.” (External partner at the national level)* | *"We have been advising facilities that they need to own the data; they have to make sure that they utilize it by analyzing... If they are not meeting their targets, they need to reflect on the data and see where they are going wrong… For those [health facilities] that are performing well, we find out what they are doing, and we try to replicate to the other facilities that are not doing as well." (District Health Officer)* |
