## Appendix 3 for "Critical success factors for high routine immunization performance: A multiple case study analysis of Nepal, Senegal, and Zambia"

### **Appendix 3 – Reflexivity Statement**

1. **How does this study address local research and policy priorities?**

To address local needs, the Emory study team partnered with local research groups in Nepal, Zambia, and Senegal. This partnership ensured the study was aligned with the context and priorities of each country. Data collection tools were iterative and were created with advice and input from both in-country researchers, local stakeholders, and external technical advisors. The tailoring of the project to each country allowed for this hypothesis generating study to best reflect the main needs and priorities of the countries surveyed.

2. **How were local researchers involved in study design?**

Local researchers were key to the realization of this project. The in-country study teams helped draw our KII list of key stakeholders and past influential leaders, along with helping to secure meetings. We worked with the local teams to develop our tools, and to edit the questions or procedures as needed based on cultural and political considerations. Partners also helped to determine the best locations for our sample and were responsible for finding individuals for FGDs [1].

3. **How has funding been used to support the local research team?**

The local research teams were treated as sub-contractors on this project and were paid fairly for their time and effort on this project. We did not have any free or un-funded contributions of local researchers on behalf of the project.

4. **How are research staff who conducted data collection acknowledged?**

Research staff who conducted data collection were acknowledged as authors in their respective country case studies [2-4].

5. **Do all members of the research partnership have access to study data?**

In-country research partners have access to all KII and FGD transcripts from their respective countries.

6. **How was data used to develop analytical skills within the partnership?**

While we did not focus on capacity building during this study, the Emory research staff conducted trainings with each in-country research group. These trainings were over 7 – 14 days, and reviewed the study protocols, data collection guidelines, and general qualitative research best practices.

7. **How have research partners collaborated in interpreting study data?**

Within each country level case study, our in-country partners were actively engaged in interpretations; the study teams had weekly meetings to discuss the progress and findings of KIIs and FGDs. In addition, partners were involved in the drafting of the manuscripts – the case studies and this cross-cutting synthesis [2-4].

8. **How were research partners supported to develop writing skills?**

Our research partners collaborated on the manuscript drafts and were added as authors for their contributions. Additionally, several student graduate assistants were not American, and from LMICs; students were added as authors for their contributions.

9. **How will research products be shared to address local needs?**

Results from this study have already been disseminated to the respective leaders and stakeholders of each country; the meeting was led by the in-country PI and was attended by the Emory research team. A country-specific report, along with the manuscripts, were disseminated. We met with our TAG to disseminate results to global immunization leaders, and continue to collaborate with the in-country PIs on opportunities for additional outreach.

10. **How is the leadership, contribution and ownership of this work by LMIC researchers recognised within the authorship?**

On this cross-country synthesis, the in-country PIs are added as senior authors. For the individual case studies, all in-country researchers are included as authors [2-4].

11. **How have early career researchers across the partnership been included within the authorship team?**

In country research partners hired their own staff, and there were different levels of experience represented across the researchers. Early career researchers were included and mentored within each in-country research team, as well as within the Emory team.

12. **How has gender balance been addressed within the authorship?**

While the Emory and in-country PI’s are men, most of the research staff was comprised of women. Guidance for authorship was drafted prior to writing manuscripts, and all were given opportunities to contribute. Those who met the criteria were included as authors; individual contributions, and specific authors, are included on the country case studies [2-4]. This manuscript is the synthesis, and country-level PIs and the Emory team are included as authors.

13. **How has the project contributed to training of LMIC researchers?**

This project had 7 – 14 day trainings for each country’s research team, wherein best practices for qualitative research were taught along with the study’s aims and tools. In-country PI’s provided additional assistance and training to staff as needed, however, our collaborators were all well-trained professionals and knowledgeable in their areas of expertise.

14. **How has the project contributed to improvements in local infrastructure?**

This project was a hypothesis generating study, and therefore did not contribute to infrastructure improvement. However, we hope the findings of this study will be of use to program managers and funders in strengthening and building further immunization infrastructure that is both resilient and sustainable.

15. **What safeguarding procedures were used to protect local study participants and researchers?**

KII and FGD participants gave researchers signed, informed consent; their information was transcribed and de-identified. See our protocol and case study papers for further information [1-4]. There was only minimal risk to both participants and researchers; local PI’s were responsible for any context specific safeguarding.

1.	Bednarczyk, R.A., et al., *Exemplars in vaccine delivery protocol: a case-study-based identification and evaluation of critical factors in achieving high and sustained childhood immunisation coverage in selected low-income and lower-middle-income countries.* BMJ Open, 2022. **12**.

2.	Micek, K., et al., *Critical success factors for routine immunization performance: A case study of Zambia 2000 to 2018.* Vaccine X, 2022. **11**.

3.	Hester, K.A., et al., *Critical success factors for high routine immunization performance: A case study of Nepal.* Vaccine X, 2022. **12**(100214).

4.	Sakas, Z., et al., *Critical success factors for high routine immunization performance: A case study of Senegal.* medRxiv, 2022: p. 2022.01.25.22269847.
